# Patterns of Health Insurance Coverage, Determinants, and User Satisfaction in Eight Sub-Saharan African Countries: Implications for Health Financing Policy and Equity

**DOI:** 10.64898/2026.01.02.26343335

**Authors:** Lizah Nyawira, Yvonne Opanga, Boniface Mbuthia, Lazarus Odeny, Richard Kiplimo, Saida Kassim, Mable Jerop, Moreen Mwenda, Omar Kopi, Jocine Ogoya, Charles Kithinji, Jane Sydney Jabilo, Norah Mwase, Samuel Muhula Opondo

## Abstract

**Introduction:** Effective risk pooling plays a critical role in advancing universal health coverage by improving financial protection and access to healthcare. However, in sub-Saharan Africa, insurance coverage remains low, and satisfaction with existing schemes is inconsistent, raising concerns about their effectiveness. This study examines the factors influencing health insurance coverage and satisfaction across eight countries in sub-Saharan Africa

**Materials and Methods:** Using household survey data, the levels and predictors of health insurance coverage as well as satisfaction with health insurance mechanisms were examined. The predictors of health insurance coverage, satisfaction with health insurance mechanisms and health care providers were modeled using a combination of bivariate and multivariate analyses.

**Results:** Overall, 24% of households included in the study reported having some form of health insurance, with wide variation across countries. Ethiopia recorded the highest coverage (55%), followed by Kenya (23%) and Senegal (17%), while the lowest levels were observed in South Sudan (7.7%) and Zambia (12%). Insurance coverage was slightly higher among men (28%) than women (22%), and uptake increased with age, education, and household income, with insured households reporting lower out-of-pocket expenditures. Multivariable analysis showed that country, age, household size, and education significantly predicted insurance enrollment; compared to Ethiopia, all other countries had substantially lower odds of coverage (aOR 0.06–0.27, p<0.001), while older age, larger households, and secondary education increased the likelihood of being insured. Insurance mechanisms varied by context, with community-based schemes dominant in Ethiopia and Uganda, social/national insurance common in Kenya, Tanzania, and Zambia, and private insurance more prevalent in Kenya, Ethiopia, and Uganda. Among the uninsured (70%), key barriers included high premium costs (38%), lack of awareness (35%), and perceived lack of value (15%), with affordability concerns more common among women. Satisfaction levels were moderate with 43% satisfied and 11% dissatisfied with their insurance, while satisfaction with healthcare providers was highest in Tanzania and Uganda, which also showed the strongest adjusted odds of satisfaction (aOR 6.60–11.85 and 8.15–8.40, respectively).

**Conclusion:** Health insurance coverage in sub-Saharan Africa remains low and uneven, reflecting deep socioeconomic and geographic disparities. Education, age, and household size significantly influence enrollment, highlighting the role of awareness and affordability. High premium costs and limited understanding remain major barriers to participation. Satisfaction with insurance and healthcare providers is moderate, with many respondents expressing neutrality about service quality. Strengthening health financing requires subsidies for vulnerable groups, better awareness campaigns, and integration of community schemes into national systems. Such measures are vital to promote equity, sustainability, and progress toward universal health coverage. This demands reimagining benefit design, strengthening provider networks, and leveraging strategic purchasing to drive both equity and satisfaction

## 1.0 Introduction

Health insurance is a form of risk pooling mechanism designed to protect households from catastrophic healthcare expenses while enhancing access to essential health services [1]. Contributory health insurance requires beneficiaries to pay premiums directly often through employers or self-contributions while non-contributory health insurance is fully funded by the government or donors, providing coverage without direct payments from beneficiaries[2]. The effectiveness of a health insurance mechanism is often measured by coverage levels and user satisfaction, as these factors reflect its ability to improve healthcare access, provide financial protection, and ensure service quality [3].

Global health insurance coverage remains highly uneven across regions. In Europe, most countries have achieved near-Universal Health Coverage (UHC) through social health insurance or tax-based systems, with service coverage indices averaging above 70% across the region [4–5]. In contrast, coverage in the Americas show significant disparities such that while over 90% of individuals in the United States are insured, rates remain considerably lower across Latin America and the Caribbean [6]. The World Health Organization (WHO) Western Pacific Region demonstrates moderate progress, with a UHC service index of approximately 75%, reflecting expanding access through national schemes such as those in Vietnam, Indonesia, and the Philippines [7–8]. Africa, however, continues to lag behind, with only about 17% of the population covered by any form of health insurance [4,9]. These regional disparities underscore persistent inequities in financial protection and access to essential health services globally.

In sub-Saharan Africa (SSA), health insurance coverage remains low and highly fragmented, with majority of the population still relying on out-of-pocket (OOP) payments, which expose them to catastrophic health expenditure (CHE) and limit healthcare access [10]. According to the WHO, CHE typically occurs when OOP health payments exceed 10% or 25% of household income or total expenditure [11]. In SSA, where over 40% of total health spending is financed through OOP payments, millions of families experience financial distress and impoverishment due to healthcare costs [4, 12]. Despite growing global efforts to expand financial protection and make progress towards UHC, the region faces several structural, economic, and policy-related barriers that hinder widespread health insurance adoption. OOP costs despite coverage discourage participation [13], while low public health spending leads to under-resourced insurance programs [14]. The dominance of informal employment in Africa makes it difficult to collect premiums or extend coverage beyond formal sector workers [15]. Additionally, limited public awareness, low trust in health systems, poor service quality, and widespread poverty hinder enrollment efforts [16–18]. Table 1 depicts health financing and other relevant socio-demographic indicators in countries in Africa.

**Table 1:** Health Financing Sources and Related Indicators across the surveyed countries.

| Health Financing Sources and Related Indicators |  | Population 2023* (millions) | GDP per Capita 2023* | Donor financing** (%) | Public Financing (%) | OOP payments** (%) | Health insurance coverage** (%) | GHE per capita** | GHE as % of GDP*** (%) | Catastrophic expenditure (10% of household budget)**** | Catastrophic expenditure (25% of households budget) **** |
| --- | --- | --- | --- | --- | --- | --- | --- | --- | --- | --- | --- |
| Country | Ethiopia | 128.7 | 1,272.0 | 33.9 | 32.2 | 30.5 | 2.5 | 6.84 | 0.72 | 3.5 | 0.6 |
|  | South Sudan | 11.5 | 1,080.1 | 49.8 | 15.9 | 27.4 | 4.8 | 4.24 | 0.58 | 11.7 | 2.7 |
|  | Kenya | 55.3 | 1,952.3 | 17.8 | 52.3 | 22.7 | 25 | 42.27 | 2.02 | 5.2 | 1.4 |
|  | Uganda | 48.7 | 1,002.3 | 42.9 | 17.5 | 38.6 | 2.0 | 9.83 | 0.98 | 15.3 | 3.8 |
|  | Tanzania | 66.6 | 1,224.5 | 30 | 40 | 22 | 32 | 11.25 | 0.98 | 4.3 | 0.8 |
|  | Malawi | 21.1 | 602.3 | 54.5 | 24.1 | 11.9 | 9.3 | 5.35 | 0.88 | 2.9 | 0.4 |
|  | Zambia | 20.7 | 1,330.7 | 35.8 | 49.2 | 7.1 | 6.5 | 36.28 | 2.51 | 0.3 | 0.0 |
|  | Senegal | 18.1 | 1,706.4 | 18.4 | 21 | 47.3 | 5.3 | 14.05 | 0.9 | 6.9 | 1.3 |

Sources: \*- *World Bank Indicators*, \*\*-*WHO Health Financing Indicators (2022)*

Dissatisfaction of insured individuals with the health insurance schemes arises due to limited provider choices, poor service quality, delays in reimbursements, and gaps in benefit packages [19]. Willingness to pay (WTP) for the health insurance schemes has remained moderate and has been shown to be influenced by factors such as socio-demographic characteristics, income and economic considerations, information availability and sources, illness-related expenses, health service factors, social capital and solidarity [20]. Further, retention within the schemes is particularly influenced by perceived quality of care, provider responsiveness, and ease of claims processing [21].

While some countries have attempted to implement mandatory health insurance (e.g., Ghana’s National Health Insurance Scheme) and Kenya’s new Social Health Insurance Fund (SHIF), sustainability challenges persist due to low enrollment rates, inadequate premium collection, and inefficiencies within the health insurance agencies [22,23]. Understanding the factors that influence both coverage and satisfaction is essential for improving the design and implementation of health insurance mechanisms in the region. These factors are closely linked to Strategic Health Purchasing (SHP), which aims to enhance efficiency, quality, and equity by aligning financial resources with provider performance. SHP involves defining service entitlements, selectively contracting high-performing providers, and monitoring performance to achieve health system goals - access to quality services and financial risk protection [22, 24]. By identifying barriers to insurance uptake and retention, purchasing agencies can undertake evidence-based reforms in benefit package design, provider contracting and provider payment mechanisms.

To examine the key predictors of health insurance coverage and satisfaction with health insurance schemes and health care providers, this study employs a regression model analysis using household survey data from eight Sub-Saharan African countries (Ethiopia, South Sudan, Kenya, Uganda, Tanzania, Malawi, Zambia and Senegal) where Amref Health Africa works. This analytical approach allows for a systematic evaluation of socio-economic, demographic, and institutional factors that shape enrollment decisions and user experiences. Key variables examined included: socio-economic factors such as age, gender, education level, household size, number of dependents; and health insurance factors such as type of coverage and premiums, satisfaction with health insurance mechanisms and health provider factors (satisfaction with health services). The study aims to provide insights into existing gaps within the region, challenges, and opportunities to support policymakers and development partners in designing effective risk pooling mechanisms and enhance financial protection and equitable access to quality health care.

## Methods

### Study design

The study design adopted has been documented in another publication from this work [25]. We utilized a cross-sectional study design incorporating a quantitative approach to assess health insurance coverage levels and satisfaction with both health insurance mechanisms and the health care providers in eight study countries

### Study sites

The study was conducted in eight African countries including Ethiopia, South Sudan, Kenya, Uganda, Tanzania, Malawi, Zambia, and Senegal where Amref Health Africa implements health programs (Fig 1). These countries were purposively selected to represent a diverse cross-section of SSA, encompassing eastern, western, and southern regions. The selection was informed not only by Amref’s programmatic presence but also by the diversity in health system structures, health insurance coverage models, and progress toward UHC. In Ethiopia, the study was conducted in Oromia, Amhara, Afar, South Ethiopia, and Addis Ababa. In Kenya, eight counties were targeted including Nairobi, Narok, Vihiga, Siaya, Nyeri, West Pokot, Samburu, and Tharaka Nithi to reflect urban, rural, arid, and semi-arid contexts. In Uganda, data were collected from Iganga, Mayuge, Bugiri, Namayinga, and Pader districts. The Tanzania sites covered seven mainland regions including Singida, Mara, Songwe, Lindi, Morogoro, Tanga, and Tabora and two in Zanzibar Kaskazini Unguja and Kaskazini Pemba. In Malawi, the study was conducted in Mchinji, Chitipa, Karonga, Salima, Mangochi, and Chikwawa districts, while in Zambia, data collection occurred in Kabwe (Central Province), Kitwe (Copperbelt), Mwense (Luapula), and Sinda (Eastern). In South Sudan, research covered Warrap (Twic East, Tonj South), Western Bahr el Ghazal (Wau), Eastern Equatoria (Kapoeta South), Central Equatoria (Juba, Kajokeji), and Western Equatoria (Yambio, Maridi, Nzara). Finally, in Senegal, the study was undertaken in Kolda, Sédhiou, Goudomp, Matam, and Guédiawaye districts.

**Figure 1:**
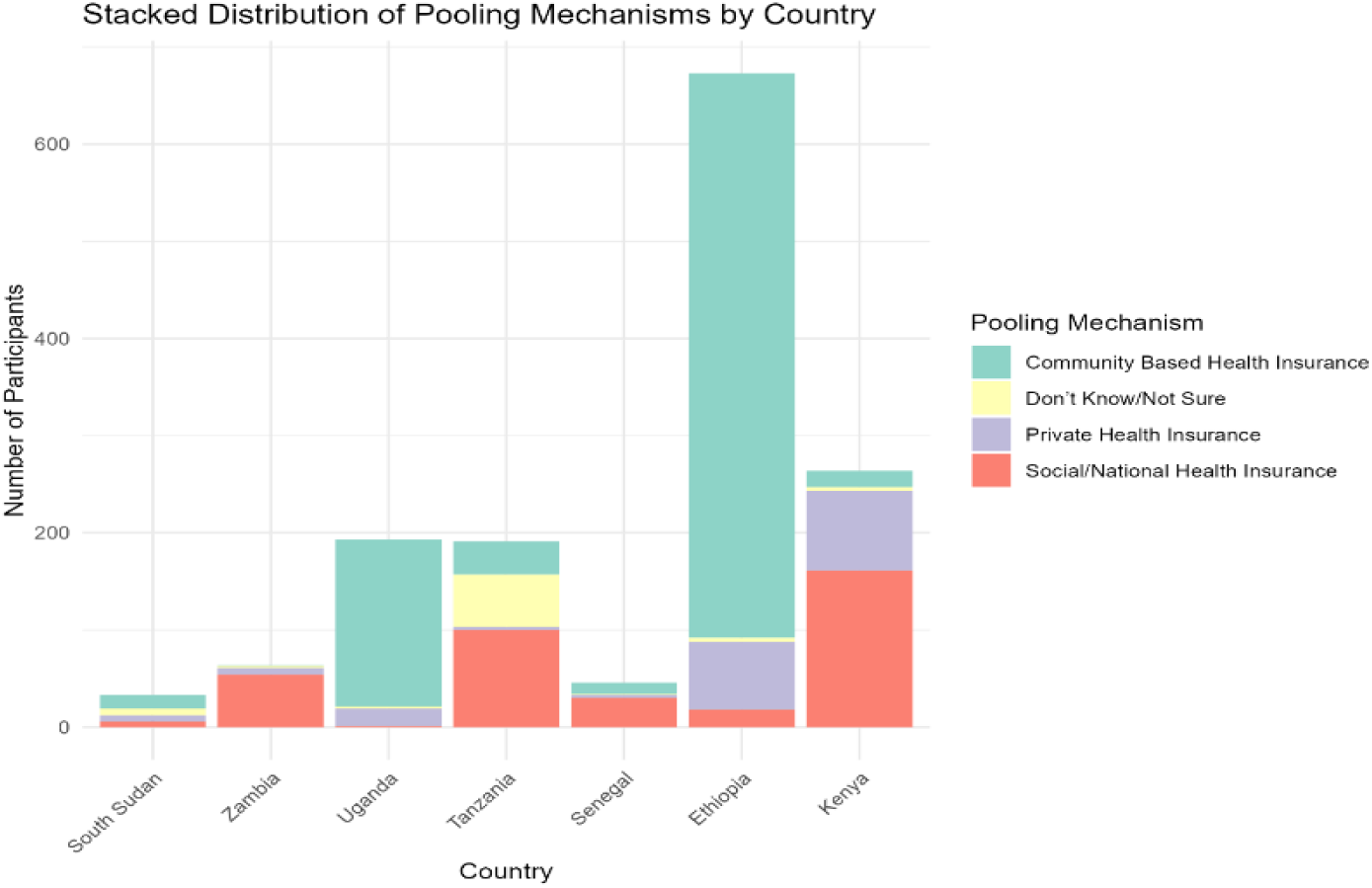
**Health insurance type by country**

Across these countries, health insurance remains a critical yet unevenly developed component of health financing, with a significant proportion of the population continuing to rely on OOP payments to access healthcare. The study aimed to capture a wide range of financial protection contexts by including countries with varied experiences in implementing national and community-based health insurance schemes. This multi-country approach enhanced understanding of different health financing contexts and levels of insurance coverage and enabled cross-country comparisons to identify region-specific determinants that shape the effectiveness of health financing. The inclusion of these countries was, therefore, a deliberate and strategic decision to ensure that the study findings are contextually grounded, reflective of the diversity within SSA, and capable of informing actionable policies and programs to strengthen health systems and expand equitable health insurance coverage across the continent.

### Study Population and Sampling

The study population comprised household heads or primary caregivers who had resided in the selected regions for at least six months prior to data collection. The target population included both men and women, categorized as youth (15–35 years) and adults (over 35 years). Eligible respondents were required to be knowledgeable about household health-seeking behaviors, healthcare expenditures, and participation in any health insurance schemes where applicable. Households were systematically sampled to ensure representation across urban, rural, arid, and semi-arid settings.

The sample size for each category was computed using parameters that accounted for the required sample size (n), average cluster size at each stage (M₁, M₂ᵢⱼ), proportions of units to be sampled (π₁, π₂ᵢⱼ), confidence interval (Z), margin of error (MOE), and design effect (DEFF). This approach ensured representativeness and adaptability to country-specific contexts. Sampling units included health facilities, health workers, clients, and households, based on a sampling framework developed by Amref Health Africa Headquarters, with country teams permitted to adjust sample sizes in response to contextual realities such as population density, accessibility, and programmatic reach.

An allowable deviation of ±10% was considered acceptable to account for non-responses and logistical constraints during fieldwork. Overall, 7,243 households and 11,994 clients were surveyed across the eight countries as indicated in Table 2.

**Table 2:** Sample Size Distribution.

| Sample Type | Ethiopia | Kenya | Tanzania | Malawi | South Sudan | Uganda | Senegal | Zambia | Total |
| --- | --- | --- | --- | --- | --- | --- | --- | --- | --- |
| **Respondents** | 2,133 | 1,279 | 1,214 | 1,010 | 532 | 3,817 | 799 | 1,210 | 11,994 |
| **Households** | 1,366 | 1,212 | 1,305 | 229 | 508 | 1,724 | 291 | 608 | 7,243 |

Although most countries achieved their target samples within the acceptable deviation range, Kenya, Malawi, and Senegal recorded slightly lower household response rates. Nevertheless, the achieved sample sizes were adequate to support robust cross-country comparisons and analysis of health financing dynamics, insurance coverage, and household-level financial protection across diverse Sub-Saharan African contexts.

### Data Analysis

All data were analyzed using R version 4.5.0 from a cross-sectional survey of 7,243 households across eight SSA countries. Descriptive statistics summarized the study population, presenting continuous variables as medians with interquartile ranges and categorical variables as frequencies and percentages. Group differences were examined using Pearson’s chi-squared and Wilcoxon rank-sum tests. Bivariate associations between health insurance coverage, satisfaction outcomes, and respondent characteristics were explored using cross-tabulations and univariable logistic regressions. Multivariable logistic regression models were then applied to adjust for confounders, with variables selected based on theoretical importance and statistical significance. Results were expressed as adjusted odds ratios (AORs) with 95% confidence intervals and *p*-values, and diagnostic checks were performed for model fit and multicollinearity. Key findings were visualized using forest plots, bar charts for satisfaction levels, and stacked bar charts showing pooling mechanism distributions, all created using the ggplot2 package in R.

### Ethical considerations

The research protocols were reviewed and approved by local ethics committees in five out of the eight countries of data collection. Ethiopia and Kenya were approved by the Amref Ethics and Scientific Review Committee (ESRC), ESRC P1580/2023 on 21 November 2023. Tanzania was approved by the National Institute for Medical Research, NIMR/HQ/R.8a/Vol.IX/4440 on 27 October 2023 and by the Zanzibar Health Research Institute, ZAHREC/04/PR/NOV/2023/35 on 17 November 2023. Zambia was approved by Eres Converge, an independent ethics board, 2023-Nov-001 on 28 November 2023 and the National Health Research Authority (NHRA) on 27 November 2023. Uganda was approved by the Uganda Christian University Review and Ethics Committee, UCUREC-2023-699 on 7 November 2023. South Sudan, Malawi, and West Africa received a waiver for a full review of their proposals from Ethics Boards because, being evaluations, they did not meet standards to undergo ethical approval according to the country standards. Waiver letter from Malawi is attached for reference.

## 3. RESULTS

### 3.1 Socio-demographic characteristics

A total of 7,243 households were surveyed across the eight study countries, with varying sample sizes. The median age of household respondents was 38 years (IQR: 15–97), and the majority were female (64%). Most respondents were household heads (71%), while the remaining were spouses or children. Educational attainment varied, with 20% of respondents having no formal education, 40% completing primary education, 27% attaining secondary education, and only 11% having tertiary or university-level education.

Household income sources were diverse, with agriculture being the most common (48%), followed by small businesses (27%), casual labor (11%), and formal employment (14%). Fishing, livestock farming, and service-based businesses accounted for smaller proportions. The median household size was five members (IQR: 4–7). About 36% of households had at least one skilled member, while 12% and 10% reported having a pregnant woman and a differently abled household member respectively at the time of the survey. Table 3 highlights the characteristics of the households.

**Table 3:** Characteristics of Households.

| Household | Characteristics | Frequency(n = 7,243) | Percentage (%) |
| --- | --- | --- | --- |
| Country | Ethiopia | 1,366 | 19% |
|  | Kenya | 1,212 | 17% |
|  | Malawi | 229 | 3.2% |
|  | Senegal | 291 | 4.0% |
|  | South Sudan | 508 | 7.0% |
|  | Tanzania | 1,305 | 18% |
|  | Uganda | 1,723 | 24% |
|  | Zambia | 609 | 8.4% |
| Respondent Gender | Gender | Frequency (n) | Percentage (%) |
|  | Female | 4,634 | 64% |
|  | Male | 2,609 | 36% |
| Age Median | Respondent Age<br>(Median, IQR) | 38 (30, 50) |  |
| Respondent Education Level | Educational Level | Frequency (n) | Percentage (%) |
|  | None (never been to school) | 1,425 | 20% |
|  | Primary | 2,918 | 40% |
|  | Secondary | 1,964 | 27% |
|  | Tertiary/university | 816 | 11% |
|  | Vocational | 120 | 1.7% |
| **Respondent is Head of Household?** | **Head of Household** | **Frequency (n)** | **Percentage (%)** |
|  | Yes | 5,172 | 71% |
|  | No | 2,071 | 29% |
| **Household Size** | **Total household Size (Median, IQR)** | 5 (4, 7) |  |
| **Income source** | **Income source** | **Frequency (n)** | **Percentage (%)** |
|  | Agriculture | 3,512 | 48% |
|  | Fishing | 173 | 2.4% |
|  | Livestock Farming | 702 | 9.7% |
|  | Rental Houses | 307 | 4.2% |
|  | Handicrafts/Artisanal Work | 218 | 3.0% |
|  | Small Business | 1,972 | 27% |
|  | Online Business | 55 | 0.8% |
|  | Service-Based Business | 253 | 3.5% |
|  | Employment | 992 | 14% |
|  | Remittances | 143 | 2.0% |
|  | Casual Labour | 769 | 11% |
|  | Other | 553 | 8.6% |
| **Abled Members** | **Any differently abled members? (Y)** | 742 | 10% |
| **Pregnant Women in the Household** |  | **Frequency (n)** | **Percentage (%)** |
|  | Yes | 867 | 12% |
|  | No | 6,376 | 88% |

### 3.2 Health insurance coverage levels by socio-demographic characteristics of study participants

Overall, only 1,573 (24%) of surveyed households had some form of health insurance coverage, with significant disparities across countries (p <0.001). Ethiopia had the highest coverage (55%), followed by Kenya (23%), Senegal (17%), Tanzania (16%), Uganda (13%), Zambia (12%), and South Sudan (7.7%). Only one respondent from Malawi reported health insurance coverage and therefore Malawi was excluded from further analysis.

Insurance coverage was higher among men (28%) compared to women (22%). The median age of insured individuals was slightly higher at 40 years, compared to 38 years among the uninsured. Household size did not show significant differences between insured and uninsured households (median: 5 members for both groups). However, uninsured households had a lower median number of dependents (3 vs. 5; p <0.001).

Education was significantly associated with health insurance enrollment (p < 0.001). Among those with primary education, 20% were insured, increasing to 24% for those with secondary education and 35% for tertiary graduates. Surprisingly, 28% of individuals with no formal education had insurance, a higher rate than those with primary, secondary, or vocational training.

Economic factors also played a role in insurance enrollment. Uninsured households reported a lower median income of $26 (IQR: $8–$77), compared to $53 (IQR: $18–$118) for insured households. However, uninsured households also reported significantly higher out-of-pocket health expenditures (median: $5; IQR: $2–$14) compared to insured households (median: $4; IQR: $3–$13). Table 4 highlights the health insurance coverage levels by socio-demographic characteristics of study participants.

**Table 4:**
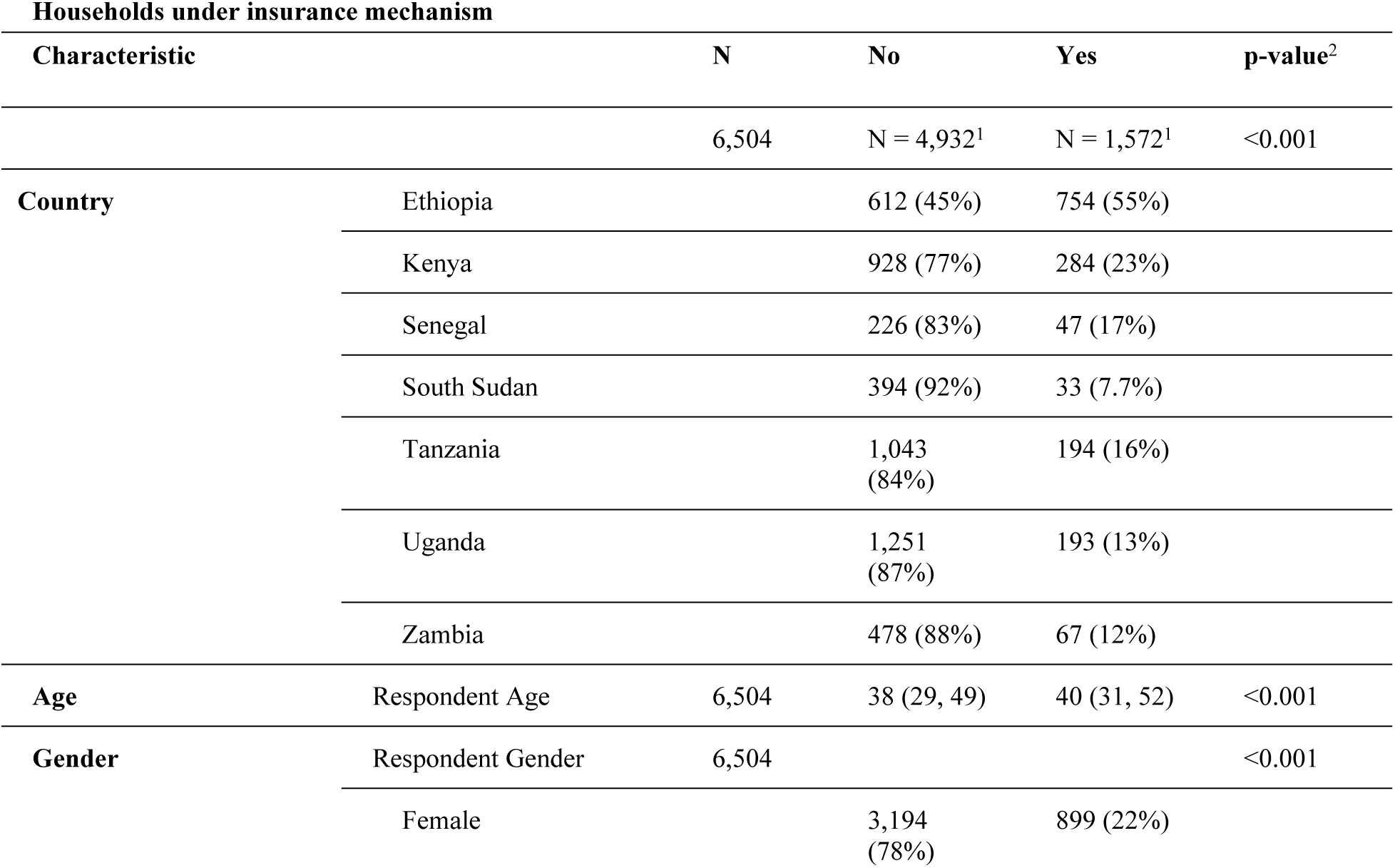
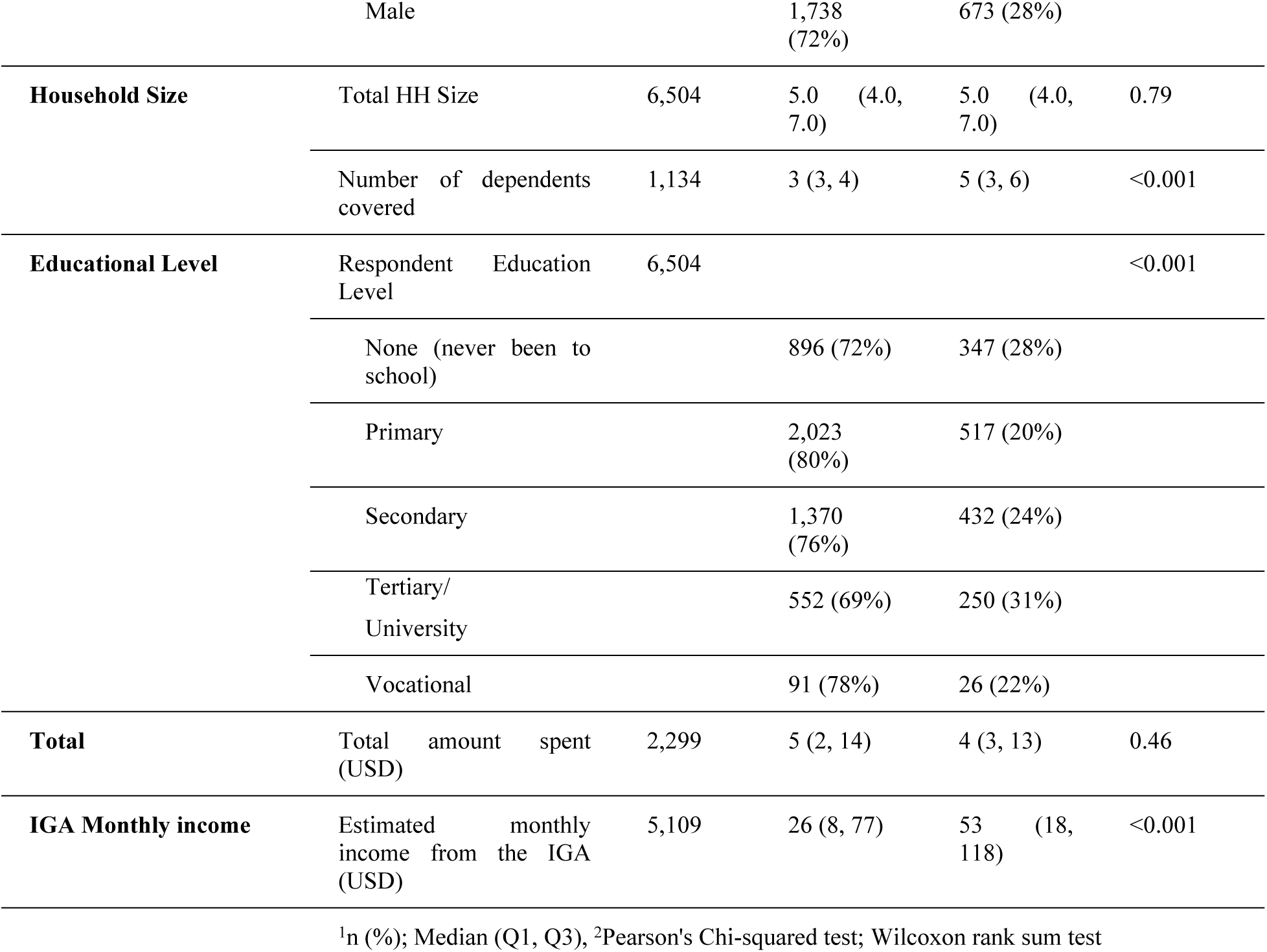
Health insurance coverage levels by socio-demographic characteristics of households – descriptive approach.

### 3.3 Predictors of health insurance coverage – analytical approach

Health insurance coverage was significantly associated with country, age, household size, and education level. Compared to Ethiopia, the odds of being insured were significantly lower in Kenya (aOR=0.26, 95% CI: 0.21–0.30), Senegal (aOR=0.11, 95% CI: 0.07–0.16), South Sudan (aOR=0.06, 95% CI: 0.04–0.09), Tanzania (aOR=0.13, 95% CI: 0.10–0.16), Uganda (aOR=0.11, 95% CI: 0.09–0.13), and Zambia (aOR=0.10, 95% CI: 0.07–0.13), all with p-values < 0.001.

Insurance enrollment increased with respondent age (aOR=1.02, 95% CI: 1.01–1.02, p<0.001) and household size (aOR=1.05, 95% CI: 1.03–1.07, p<0.001). Education level also influenced insurance uptake. Compared to those with no formal education, respondents who had completed secondary school had significantly higher odds of being insured (aOR=1.24, 95% CI: 1.02–1.51, p=0.033), while other education levels were not significantly associated with coverage. Gender was not a significant predictor in the adjusted model (aOR=0.98, 95% CI: 0.86–1.12, p=0.797). Table 5 presents the adjusted odds of health insurance enrollment across these variables.

**Table 5:**
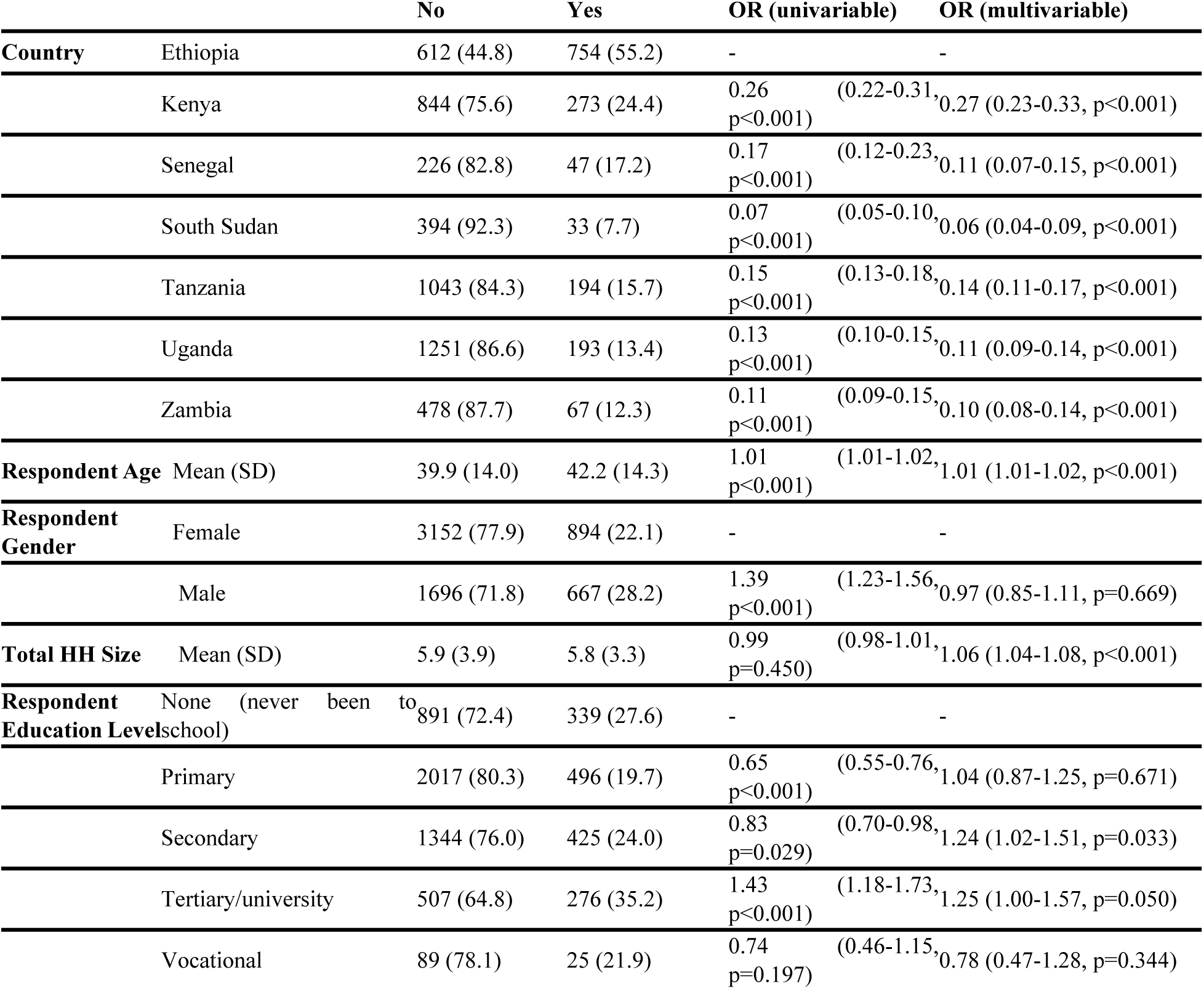
Household under insurance or pooling mechanism (The Odds of being in a Pooling Mechanism.

**Table 6:** Types of health insurance mechanisms by country.

|  |  | Country |  |  |  |  |  |  |  |
| --- | --- | --- | --- | --- | --- | --- | --- | --- | --- |
| Characteristic | N | Overall<br>N =<br>1,489 | Ethiopia<br>N = 682 | Kenya<br>N = 273 | Senegal<br>N = 47 | South<br>Sudan<br>N = 33 | Tanzania<br>N = 194 | Uganda<br>N = 193 | Zambia<br>N = 67 |
| **Type_health**<br>**insurance_mechanism** | 1,489 |  |  |  |  |  |  |  |  |
| Community-Based<br>Insurance |  | 836 | 586<br>(70%) | 17<br>(2.0%) | 12<br>(1.4%) | 14<br>(1.7%) | 34 (4.1%) | 172<br>(21%) | 1<br>(0.1%) |
| Don't Know / Not Sure |  | 74 | 4 (5.4%) | 4<br>(5.4%) | 1<br>(1.4%) | 7<br>(9.5%) | 54 (73%) | 2<br>(2.7%) | 2<br>(2.7%) |
| Private Health<br>Insurance |  | 192 | 73<br>(38%) | 82<br>(43%) | 3<br>(1.6%) | 6<br>(3.1%) | 3 (1.6%) | 18<br>(9.4%) | 7<br>(3.6%) |
| Social/National Health<br>Insurance |  | 387 | 19<br>(4.9%) | 170<br>(44%) | 31<br>(8.0%) | 6<br>(1.6%) | 103<br>(27%) | 1<br>(0.3%) | 57<br>(15%) |

<sup>1</sup>n (%)

**Table 7:**
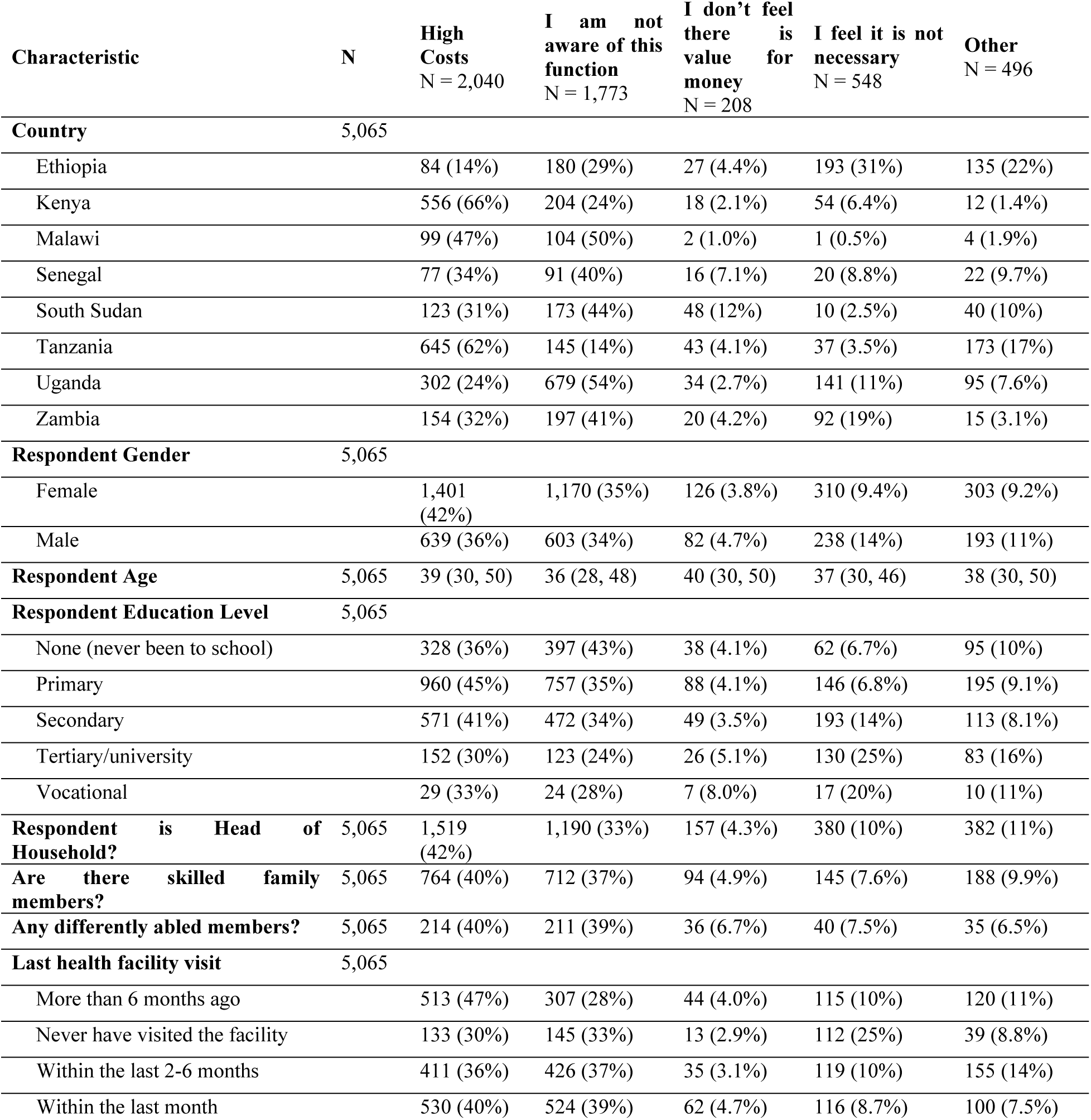
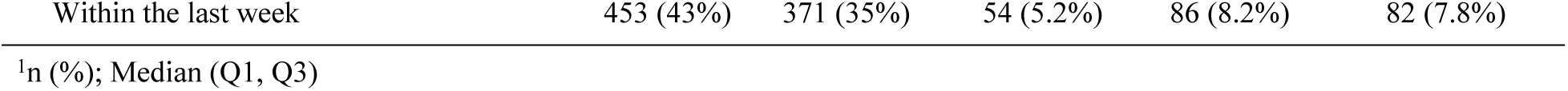
Reasons for Not Having Insurance.

### 3.4 Types of Health Insurance Mechanisms by Country

The respondents were enrolled in either national/social, community-based or private health insurance mechanisms, varying across the study countries. National or social health insurance schemes were most common in Tanzania, Senegal, Kenya, and Zambia. Community-Based Health Insurance (CBHI) schemes were dominant in Ethiopia and Uganda. Private health insurance was more prevalent in Kenya, Ethiopia, and Uganda, while Malawi and South Sudan reported low health insurance coverage across all categories. Fig 1 highlights the distribution of health insurance types by country.

### 3.5 Reasons for Non-Enrolment

Among the 5,065 uninsured respondents, 70% of the study population, the main barriers to health insurance enrollment were high premium costs (38%), lack of awareness (35%), and perceived lack of necessity or value (15%). These determinants of non-enrollment varied by country and demographic group: cost was a dominant concern in Kenya and Tanzania, while lack of awareness was more prevalent in Uganda. Financial constraints were the most frequently cited reason overall, especially among women, with 52% pointing to affordability compared to 46% of men. Meanwhile, men were more likely to question the value of insurance (39%) or claim it was unnecessary (9%). A notable portion (15%) reported being in the process of enrolling, indicating a potential for increased coverage with better follow-up.

### 3.6 Satisfaction levels with health insurance mechanism and health providers – descriptive approach

Respondents were asked about their satisfaction with both health insurance services and healthcare providers. The findings revealed a complex satisfaction landscape: Neutral responses were the most common, with 40% expressing neither satisfaction nor dissatisfaction with their insurance coverage and 44% holding a neutral opinion about healthcare provider services. Satisfied responses were slightly higher for insurance (43%) than for healthcare providers (37%). Dissatisfaction was more prevalent for healthcare providers (12%) than insurance services (11%), highlighting potential concerns with service quality, wait times, or treatment experiences. Extremely polarized opinions were rare, with only 0.9% of respondents being very dissatisfied with either insurance or provider services, while about 5.9% were very satisfied with insurance and 5.4% with providers. Fig 2 demonstrates the satisfaction spectrum for both the health insurance cover and the health providers.

**Figure 2:**
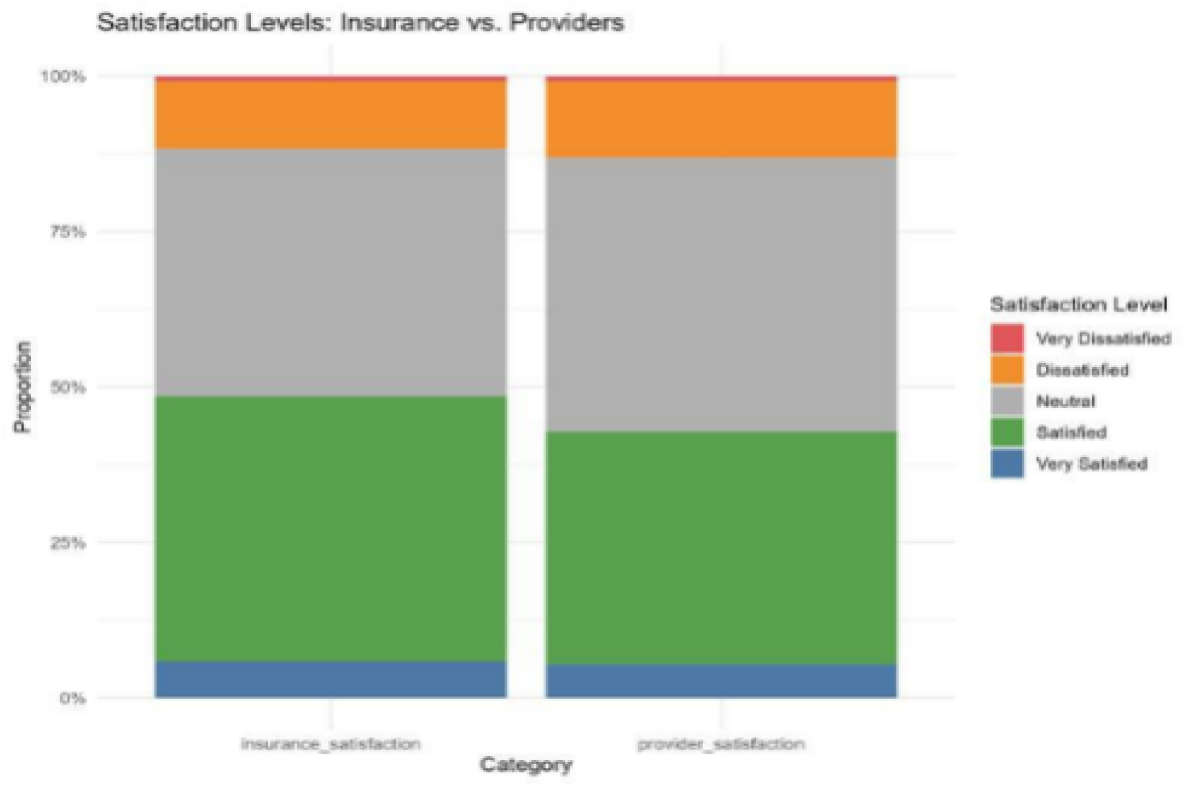
**Satisfaction spectrum for the health insurance cover and the health providers.**

### 3.7 Satisfaction with health insurance mechanism – analytical approach

Satisfaction with insurance mechanisms varied significantly across countries. Respondents from Tanzania and Uganda reported markedly higher satisfaction levels compared to Ethiopia, where 79.4% of respondents were satisfied. In Tanzania, 96.0% of respondents reported satisfaction, with both univariable (OR = 6.27, 95% CI: 2.69–18.34, p < 0.001) and multivariable models (OR = 6.60, 95% CI: 1.21–56.17, p = 0.047) showing strong and statistically significant associations. Uganda showed a similarly strong pattern, with adjusted odds of satisfaction over eleven times higher than Ethiopia (OR = 11.85, 95% CI: 1.80–243.16, p = 0.031).

Respondents in Kenya and Zambia also had elevated odds of satisfaction. While the association for Kenya was significant in the adjusted model (OR = 4.12, 95% CI: 1.22–16.66, p = 0.032), the result for Zambia, though high in magnitude, did not reach significance after adjustment (OR = 5.04, 95% CI: 0.77–105.54, p = 0.160). No significant differences were observed in satisfaction with health insurance mechanisms in Senegal or South Sudan relative to Ethiopia.

Gender, age, and household headship did not significantly influence satisfaction with health insurance mechanisms. Although male respondents had slightly lower odds of satisfaction in univariable analysis (OR = 0.80, p = 0.331), this relationship reversed after adjustment but remained non-significant (OR = 1.34, p = 0.543). Age was unrelated to satisfaction (adjusted OR = 1.00, p = 0.872), and household headship showed a non-significant trend toward lower satisfaction among heads of households (OR = 0.31, 95% CI: 0.08–1.05, p = 0.069).

The presence of skilled family members was associated with higher satisfaction in univariable analysis (OR = 1.84, 95% CI: 1.12–3.15, p = 0.021), but the association was no longer significant after adjusting for other variables (OR = 0.95, p = 0.925). Disability status and timing of the most recent health facility visit were not significantly associated with satisfaction. Those who visited a facility within the last month had lower odds of satisfaction in univariable analysis (OR = 0.36, p = 0.007), but this relationship did not persist in the multivariable model (OR = 0.70, p = 0.553).

Household decision-making power on financial matters showed mixed results. Respondents with limited or no say had lower odds of being satisfied in unadjusted models (OR = 0.33, p = 0.064), though this association was not statistically significant in adjusted analyses (OR = 0.36, p = 0.142). Sole decision-makers did not significantly differ from those in joint decision-making households. Fig 3/Table 8 depicts the predictors of satisfaction with health insurance mechanisms using regression modelling.

**Figure 3:**
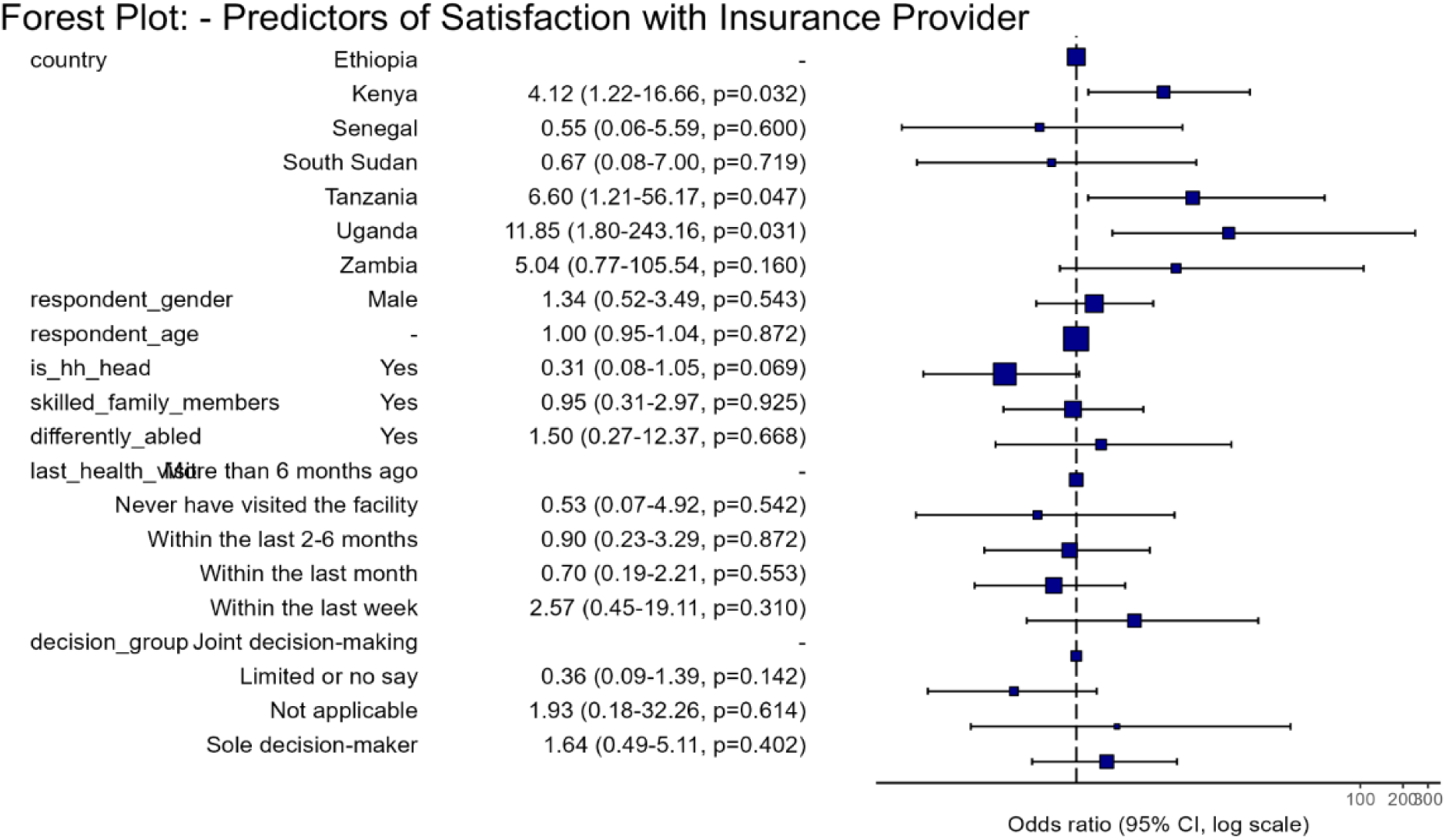
**Predictors of satisfaction with health insurance mechanism**

**Table 8:** Predictors of satisfaction with the health insurance mechanism.

| Insurance Satisfaction: |  | Dissatisfied | Satisfied | OR (univariable) | OR (multivariable) |
| --- | --- | --- | --- | --- | --- |
| Country | Ethiopia | 57 (20.6) | 220 (79.4) | - | - |
| | Kenya | 13 (12.7) | 89 (87.3) | 1.77 (0.95-3.53, $p=0.084$) | 4.12 (1.22-16.66, $p=0.032$) |
| | Senegal | 4 (14.8) | 23 (85.2) | 1.49 (0.55-5.23, $p=0.478$) | 0.55 (0.06-5.59, $p=0.600$) |
| | South Sudan | 3 (14.3) | 18 (85.7) | 1.55 (0.50-6.80, $p=0.491$) | 0.67 (0.08-7.00, $p=0.719$) |
| | Tanzania | 5 (4.0) | 121 (96.0) | 6.27 (2.69-18.34, $p<0.001$) | 6.60 (1.21-56.17, $p=0.047$) |
| | Uganda | 7 (8.3) | 77 (91.7) | 2.85 (1.33-7.09, $p=0.013$) | 11.85 (1.80-243.16, $p=0.031$) |
| | Zambia | 2 (4.8) | 40 (95.2) | 5.18 (1.53-32.41, $p=0.026$) | 5.04 (0.77-105.54, $p=0.160$) |
| Respondent_gender | Female | 48 (12.3) | 342 (87.7) | - | - |
| | Male | 43 (14.9) | 246 (85.1) | 0.80 (0.52-1.25, $p=0.331$) | 1.34 (0.52-3.49, $p=0.543$) |
| Respondent_age | Mean (SD) | 43.9 (14.1) | 44.3 (14.6) | 1.00 (0.99-1.02, $p=0.764$) | 1.00 (0.95-1.04, $p=0.872$) |
| Is_hh_head | No | 15 (10.9) | 123 (89.1) | - | - |
| | Yes | 76 (14.0) | 465 (86.0) | 0.75 (0.40-1.31, $p=0.329$) | 0.31 (0.08-1.05, $p=0.069$) |
| Skilled_family_members | No | 70 (15.6) | 379 (84.4) | - | - |
| | Yes | 21 (9.1) | 209 (90.9) | 1.84 (1.12-3.15, $p=0.021$) | 0.95 (0.31-2.97, $p=0.925$) |
| Differently_abled | No | 82 (13.3) | 535 (86.7) | - | - |
| | Yes | 9 (14.5) | 53 (85.5) | 0.90 (0.45-2.02, $p=0.787$) | 1.50 (0.27-12.37, $p=0.668$) |
| Last_health_visit | More than 6 months ago | 10 (7.7) | 120 (92.3) | - | - |
|  | Never have visited the facility | 5 (16.7) | 25 (83.3) | 0.42 (0.14-1.43, p=0.138) | 0.53 (0.07-4.92, p=0.542) |
|  | Within the last 2-6 months | 23 (13.1) | 153 (86.9) | 0.55 (0.24-1.18, p=0.138) | 0.90 (0.23-3.29, p=0.872) |
|  | Within the last month | 40 (18.7) | 174 (81.3) | 0.36 (0.17-0.73, p=0.007) | 0.70 (0.19-2.21, p=0.553) |
|  | Within the last week | 13 (10.1) | 116 (89.9) | 0.74 (0.31-1.76, p=0.501) | 2.57 (0.45-19.11, p=0.310) |
| Decision_group | Joint decision-making | 6 (10.3) | 52 (89.7) | - | - |
|  | Limited or no say | 8 (25.8) | 23 (74.2) | 0.33 (0.10-1.06, p=0.064) | 0.36 (0.09-1.39, p=0.142) |
|  | Not applicable | 2 (20.0) | 8 (80.0) | 0.46 (0.09-3.52, p=0.391) | 1.93 (0.18-32.26, p=0.614) |
|  | Sole decision-maker | 18 (12.9) | 121 (87.1) | 0.78 (0.27-1.97, p=0.611) | 1.64 (0.49-5.11, p=0.402) |

OR = Odds Ratio,

### 3.8 Satisfaction with health care providers – analytical approach

Significant differences in satisfaction with healthcare providers were observed across countries. Compared to Ethiopia, where 76.6% of respondents reported satisfaction, those in Tanzania (93.8% satisfied) and Uganda (91.0%) had notably higher odds of satisfaction. In univariable models, respondents in Tanzania had over four times the odds of being satisfied (OR = 4.62, 95% CI: 2.35–10.17, p < 0.001), and those in Uganda had more than triple the odds (OR = 3.09, 95% CI: 1.51–7.21, p = 0.004). These associations remained strong in the multivariable models: Tanzania (OR = 8.15, 95% CI: 2.04–43.95, p = 0.006) and Uganda (OR = 8.40, 95% CI: 2.01–58.64, p = 0.010). Kenya also showed elevated odds of satisfaction relative to Ethiopia (multivariable OR = 3.02, 95% CI: 0.99–10.37, p = 0.062), although this just missed conventional significance. No statistically significant differences were found for Senegal, South Sudan, or Zambia.

Sociodemographic characteristics were generally not strongly associated with satisfaction with health providers. Gender differences were minimal, with males slightly less likely to report satisfaction compared to females, though the difference was not statistically significant (multivariable OR = 0.71, 95% CI: 0.30–1.65, p = 0.424). Age was also not significantly associated with satisfaction (OR = 0.98, 95% CI: 0.95–1.02, p = 0.406). Educational attainment showed no clear pattern, and differences by education level were not statistically significant in adjusted models. For example, respondents with tertiary education had slightly higher satisfaction rates than those with no education, but the difference was not meaningful (OR = 0.81, 95% CI: 0.21–3.01, p = 0.755).

Household headship was unrelated to satisfaction, with heads of household showing similar odds to non-heads (OR = 0.62, 95% CI: 0.22–1.68, p = 0.357). Respondents from households with skilled family members had higher odds of satisfaction in the unadjusted model (OR = 1.96, 95% CI: 1.25–3.18, p = 0.004), but this association weakened after adjustment (OR = 1.97, 95% CI: 0.77–5.29, p = 0.162).

Recent healthcare utilization did not show a consistent relationship with satisfaction. Those who had visited a health facility within the past week had higher odds of satisfaction (OR = 1.44, 95% CI: 0.39–5.67, p = 0.591), but this was not statistically significant. Similarly, no meaningful differences were observed across other time intervals of last facility visit.

Decision-making power within the household, specifically regarding income use, showed a potential association with satisfaction. Sole decision-makers had higher odds of satisfaction compared to those in joint decision-making households (OR = 2.45, 95% CI: 0.97–6.25, p = 0.058), though the result narrowly missed statistical significance. Those with limited or no say, or for whom the question was not applicable, did not differ significantly from the reference group.

In summary, satisfaction with healthcare providers varied significantly by country, with particularly strong associations observed in Tanzania and Uganda. While sociodemographic factors and healthcare utilization patterns had limited influence, individual autonomy in household financial decisions may play a role in shaping provider satisfaction. Fig 4/Table 9 depicts the predictors of satisfaction with health providers.

**Figure 4:**
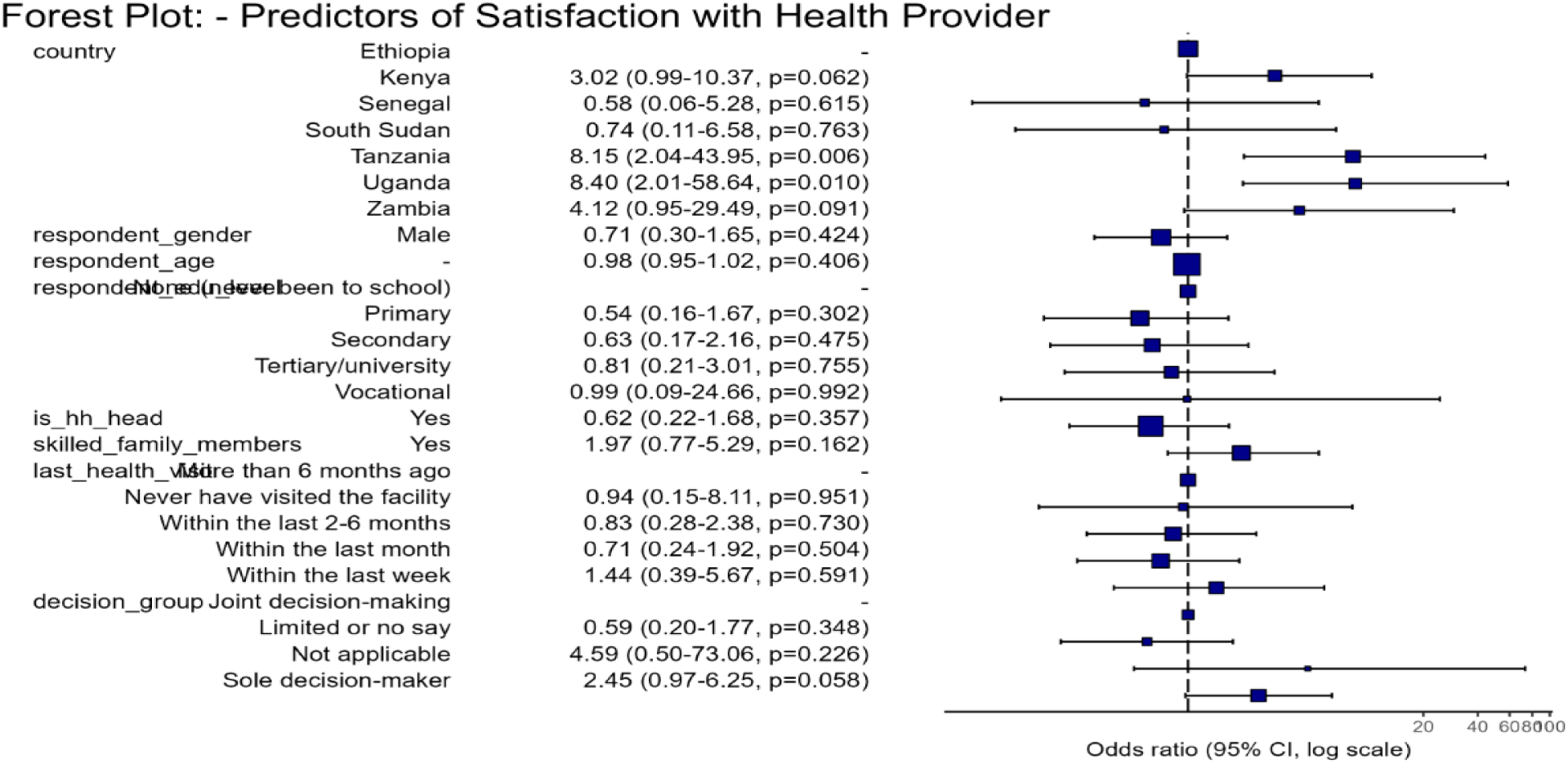
**Predictors of satisfaction with health provider**

**Table 9:**
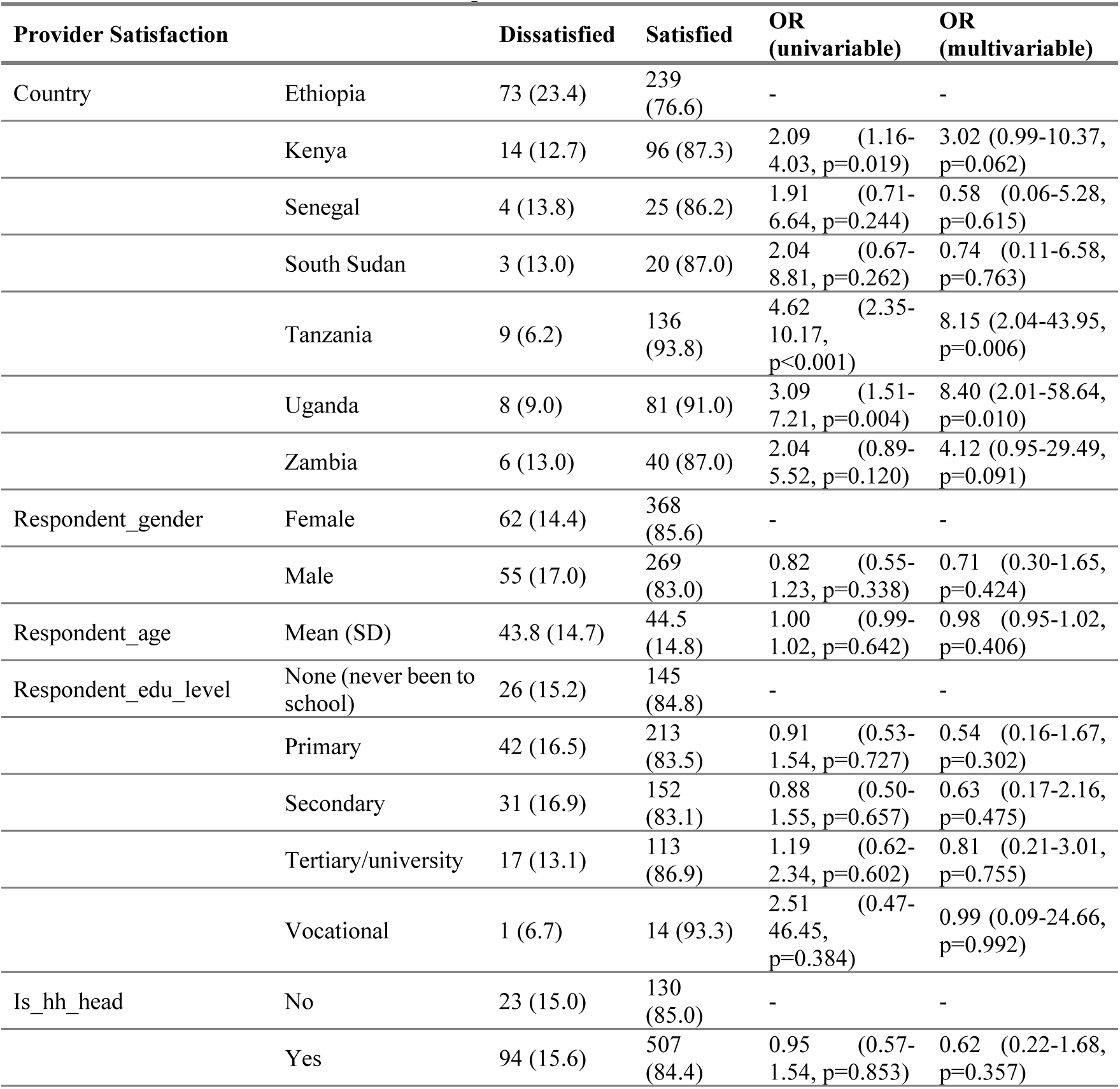
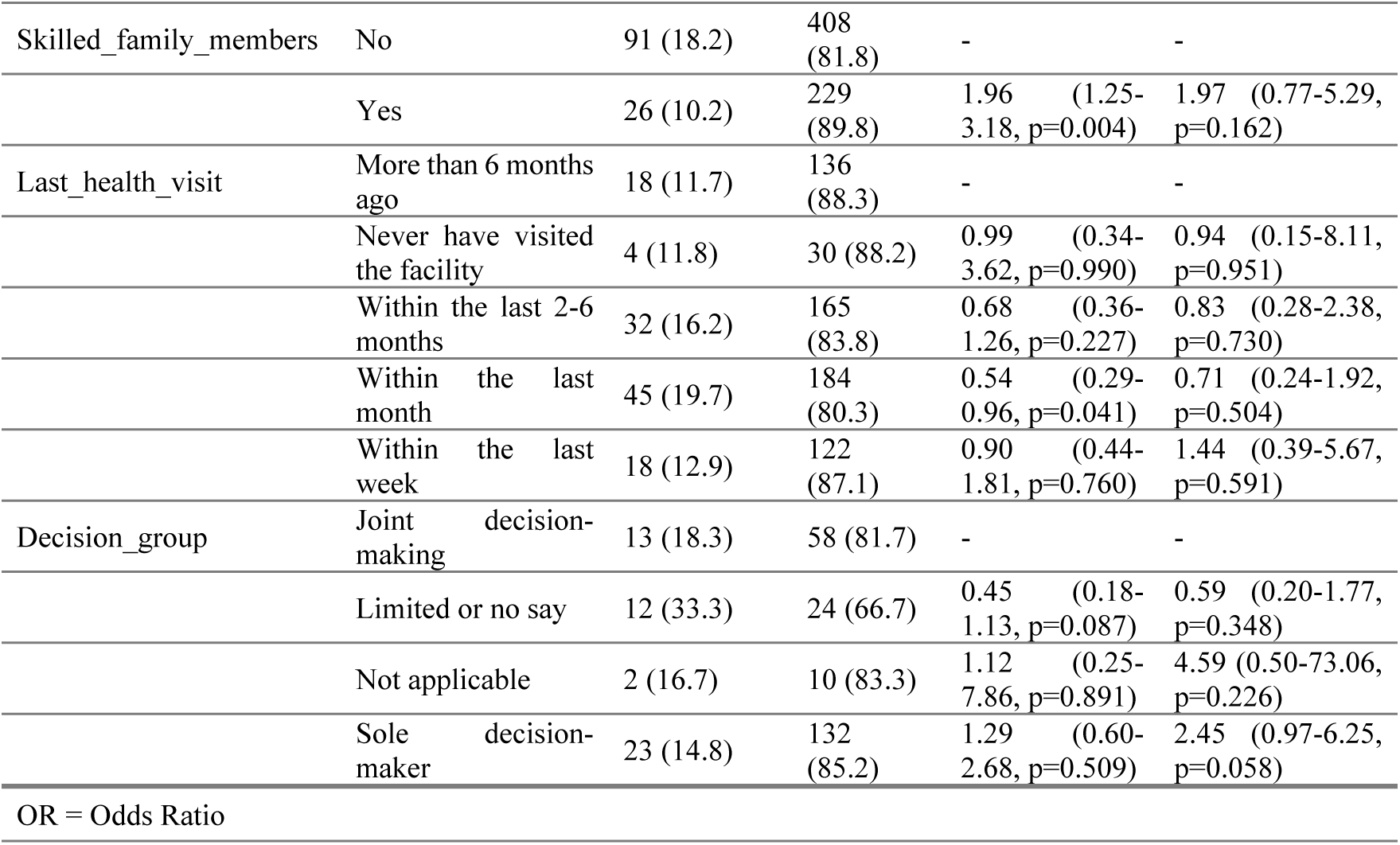
Predictors of satisfaction with health provider.

| Provider Satisfaction |  | Dissatisfied | Satisfied | OR<br>(univariable) | OR<br>(multivariable) |
| --- | --- | --- | --- | --- | --- |
| Country | Ethiopia | 73 (23.4) | 239<br>(76.6) | - | - |
| | Kenya | 14 (12.7) | 96 (87.3) | 2.09 (1.16-4.03, $p=0.019$) | 3.02 (0.99-10.37, $p=0.062$) |
| | Senegal | 4 (13.8) | 25 (86.2) | 1.91 (0.71-6.64, $p=0.244$) | 0.58 (0.06-5.28, $p=0.615$) |
| | South Sudan | 3 (13.0) | 20 (87.0) | 2.04 (0.67-8.81, $p=0.262$) | 0.74 (0.11-6.58, $p=0.763$) |
| | Tanzania | 9 (6.2) | 136<br>(93.8) | 4.62 (2.35-10.17, $p<0.001$) | 8.15 (2.04-43.95, $p=0.006$) |
| | Uganda | 8 (9.0) | 81 (91.0) | 3.09 (1.51-7.21, $p=0.004$) | 8.40 (2.01-58.64, $p=0.010$) |
| | Zambia | 6 (13.0) | 40 (87.0) | 2.04 (0.89-5.52, $p=0.120$) | 4.12 (0.95-29.49, $p=0.091$) |
| Respondent_gender | Female | 62 (14.4) | 368<br>(85.6) | - | - |
| | Male | 55 (17.0) | 269<br>(83.0) | 0.82 (0.55-1.23, $p=0.338$) | 0.71 (0.30-1.65, $p=0.424$) |
| Respondent_age | Mean (SD) | 43.8 (14.7) | 44.5<br>(14.8) | 1.00 (0.99-1.02, $p=0.642$) | 0.98 (0.95-1.02, $p=0.406$) |
| Respondent_edu_level | None (never been to school) | 26 (15.2) | 145<br>(84.8) | - | - |
| | Primary | 42 (16.5) | 213<br>(83.5) | 0.91 (0.53-1.54, $p=0.727$) | 0.54 (0.16-1.67, $p=0.302$) |
| | Secondary | 31 (16.9) | 152<br>(83.1) | 0.88 (0.50-1.55, $p=0.657$) | 0.63 (0.17-2.16, $p=0.475$) |
| | Tertiary/university | 17 (13.1) | 113<br>(86.9) | 1.19 (0.62-2.34, $p=0.602$) | 0.81 (0.21-3.01, $p=0.755$) |
| | Vocational | 1 (6.7) | 14 (93.3) | 2.51 (0.47-46.45, $p=0.384$) | 0.99 (0.09-24.66, $p=0.992$) |
| Is_hh_head | No | 23 (15.0) | 130<br>(85.0) | - | - |
| | Yes | 94 (15.6) | 507<br>(84.4) | 0.95 (0.57-1.54, $p=0.853$) | 0.62 (0.22-1.68, $p=0.357$) |
| Skilled_family_members | No | 91 (18.2) | 408 (81.8) | - | - |
|  | Yes | 26 (10.2) | 229 (89.8) | 1.96 (1.25-3.18, p=0.004) | 1.97 (0.77-5.29, p=0.162) |
| Last_health_visit | More than 6 months ago | 18 (11.7) | 136 (88.3) | - | - |
|  | Never have visited the facility | 4 (11.8) | 30 (88.2) | 0.99 (0.34-3.62, p=0.990) | 0.94 (0.15-8.11, p=0.951) |
|  | Within the last 2-6 months | 32 (16.2) | 165 (83.8) | 0.68 (0.36-1.26, p=0.227) | 0.83 (0.28-2.38, p=0.730) |
|  | Within the last month | 45 (19.7) | 184 (80.3) | 0.54 (0.29-0.96, p=0.041) | 0.71 (0.24-1.92, p=0.504) |
|  | Within the last week | 18 (12.9) | 122 (87.1) | 0.90 (0.44-1.81, p=0.760) | 1.44 (0.39-5.67, p=0.591) |
| Decision_group | Joint decision-making | 13 (18.3) | 58 (81.7) | - | - |
|  | Limited or no say | 12 (33.3) | 24 (66.7) | 0.45 (0.18-1.13, p=0.087) | 0.59 (0.20-1.77, p=0.348) |
|  | Not applicable | 2 (16.7) | 10 (83.3) | 1.12 (0.25-7.86, p=0.891) | 4.59 (0.50-73.06, p=0.226) |
|  | Sole decision-maker | 23 (14.8) | 132 (85.2) | 1.29 (0.60-2.68, p=0.509) | 2.45 (0.97-6.25, p=0.058) |

OR = Odds Ratio

## Discussion

This study explored predictors of health insurance coverage and satisfaction with both insurance mechanisms and healthcare providers across eight SSA countries - Ethiopia, South Sudan, Kenya, Uganda, Tanzania, Malawi, Zambia, and Senegal. Findings reveal substantial disparities in health insurance coverage, factors driving non-enrollment, and heterogeneous satisfaction levels and determinants across countries.

Only 24% of surveyed households reported having health insurance, with coverage varying markedly by country. Adjusted odds ratios (aORs) highlight that South Sudan, Kenya, Senegal, Tanzania, Uganda and Zambia had significantly lower odds of coverage relative to Ethiopia, which served as the baseline. Ethiopia’s comparatively higher rates may reflect more robust policies and support for CBHI. Despite literature favoring mandatory, unified insurance schemes for broader coverage [26,27], our study found that Ethiopia, dominated by fragmented CBHI schemes, led in health insurance enrollment. Similarly, in Rwanda the government-backed CBHI has achieved over 80% coverage [28,29]. Literature has highlighted the difficulty many governments face in implementing mandatory contributory schemes in a region where over 80% of the population works in the informal sector, often lacking stable income and formal channels to access insurance [30].

Socio-demographic factors including age, household size and education levels were significant predictors of health insurance uptake. First, insurance enrollment increased with respondent age. Health insurance enrolment is likely to increase with age due to rising health risks, greater need for financial protection, and more frequent healthcare use. Older adults often have more stable incomes or pensions, enabling them to afford premiums [31]. Similarly, several studies have highlighted that older individuals are more likely to enroll to health insurance schemes compared to their younger counterparts [31–34]. This could be influenced by their greater healthcare needs, decision-making power, and exposure to health education and outreach programs. Other factors such as gender did not influence health insurance enrolment which contrasts with studies that have shown that where women hold financial autonomy, enrollment may be higher [35–37].

Second, insured households had more dependents (median=5) compared to the uninsured (median=3; p=0.024). There is a likelihood that larger households have a greater exposure to financial risk from medical expenses, which increases the incentive to seek insurance as a protective measure. The presence of more dependents, especially children and the elderly, elevates healthcare needs and the probability of costly health events. Research conducted in Rwanda showed that such households are more vulnerable to health-related financial shocks, making health insurance crucial [37]. Additionally, targeted sensitization efforts by community-based and government programs often focus on these vulnerable groups, enhancing awareness and uptake. Research shows that households with more children are more likely to enroll due to increased interaction with health promotion initiatives. Thus, both perceived risk and strategic outreach may contribute to higher insurance uptake among larger households [33].

Third, insurance coverage increased with educational attainment - 20% for primary education, 24% for secondary, and 35% for tertiary. Interestingly, 28% of individuals without formal education had insurance, exceeding those with primary or vocational training. This counter-intuitive finding may reflect targeted outreach or support mechanisms for indigent populations. Overall, education likely enhances awareness, income potential, and system engagement

Finally, uninsured households had lower median incomes, reinforcing findings from Ghana [33] that link higher income to greater health insurance uptake. Further, household decision-making dynamics influenced non-enrollment to health insurance schemes. These findings highlight the need for tailored interventions that address not only financial barriers but also other social determinants of health.

Satisfaction with insurance schemes generally exceeded satisfaction with healthcare providers, though both varied significantly by country. Tanzania, Uganda, Kenya, and Zambia had higher satisfaction with insurance mechanisms compared to Ethiopia, whereas Senegal and South Sudan showed greater dissatisfaction. These trends may reflect differing levels of awareness and communication around entitlements. Regarding health provider satisfaction, Tanzania, Uganda, and Kenya again outperformed Ethiopia. This is reflective of perceived lower quality of health care service provision in countries such as Senegal, Zambia and South Sudan. Insurance expansion without parallel improvements in service delivery may not yield meaningful health gains. Studies from Nigeria [38] and Kenya [32] highlight similar dissatisfaction despite insurance coverage. Improving satisfaction with health providers requires tying payments to performance and service quality. Strategic purchasing mechanisms - such as performance-based financing - have shown success in Rwanda [39] and Cameroon [40] linking funding to outcomes and incentivizing provider responsiveness.

This study faced several limitations. In Ethiopia, coverage levels reported in the survey (over 50%) diverged from national statistics (below 2%), likely due to sampling from program-intensive areas. Additionally, data comparability across countries was constrained by variable definitions of insurance, exclusion of conflict-affected zones, and restricted field access, such as flood-affected regions in Kenya and conflict zones in Ethiopia.

## Conclusions

The findings of this study emphasize the urgent need for targeted policy interventions to expand health insurance coverage and improve satisfaction levels across SSA. While financial protection through health insurance is crucial, it must be accompanied by service quality improvements to enhance trust and uptake. Governments should prioritize affordable premium models, increase awareness campaigns, and strengthen service entitlements and provider payment mechanisms to ensure that insured individuals receive timely, high-quality care. Future research should conduct in-depth in country analysis on the factors contributing to low satisfaction levels of health insurance mechanisms and health providers.

## Data Availability

Data is available and will be provided upon request

## Acknowledgements

We thank the Amref Health Africa leadership and management for their invaluable support in making this study a success. Your approval and financial backing were instrumental in making this study a success. We also extend our gratitude to the country teams for their dedication and commitment in ensuring seamless implementation of the study. Your efforts were key to the successful completion of this study. Sincere thank you to all the study participants from the various countries who contributed their time generously to provide valuable insights and responses. Your input was essential to the success of this study. Lastly, we also appreciate the research assistants for their patience and expertise in guiding participants through the survey to enable us to have high quality data that has informed the writing of this paper

## References

1. WHO. Sustainable health financing, universal coverage and social health insurance. (2005).

2. Okungu, V., Chuma, J., Mulupi, S. & McIntyre, D. Extending coverage to informal sector populations in Kenya: Design preferences and implications for financing policy. BMC Health Serv Res 18, (2018).

3. Abu-Salim, T., Onyia, O. P., Harrison, T. & Lindsay, V. Effects of perceived cost, service quality, and customer satisfaction on health insurance service continuance. Journal of Financial Services Marketing 22, 173–186 (2017).

4. World Health Organization (2022). Global Health Expenditure Database (GHED). https://apps.who.int/nha/database

5. Paint maps (2024). UHC Service Coverage Index on World Map. Available at: https://paintmaps.com/statistics/1210/UHC-service-coverage-index-on-world-map

6. U.S. Census Bureau (2025). Health Insurance Coverage in the United States: 2024. Washington, DC: U.S. Department of Commerce. Available at: https://www.census.gov/library/publications/2025/demo/p60-288.html

7. ASEAN Magazine (2023). Universal Health Coverage in ASEAN. Association of Southeast Asian Nations. Available at: https://theaseanmagazine.asean.org/article/universal-health-coverage-in-asean

8. World Health Organization (2019). Tracking Universal Health Coverage: 2019 Global Monitoring Report. Geneva: WHO.

9. WhoOwnsAfrica (2024). Does Health Insurance in Africa Really Matter? Available at: https://www.whoownsafrica.com/health/does-health-insurance-in-africa-really-matter-10-reasons/

10. McIntyre, D., Obse, A. G., Barasa, E. W. & Ataguba, J. E. Challenges in Financing Universal Health Coverage in Sub-Saharan Africa. in Oxford Research Encyclopedia of Economics and Finance (Oxford University Press, 2018). doi:10.1093/acrefore/9780190625979.013.28.

11. World Health Organization (2021). Global Monitoring Report on Financial Protection in Health 2021. Geneva: WHO.

12. World Bank (2023). Tracking Universal Health Coverage: 2023 Global Monitoring Report. Washington, DC: World Bank.

13. Akweongo, P., Aikins, M., Wyss, K., Salari, P. & Tediosi, F. Insured clients out-of-pocket payments for health care under the national health insurance scheme in Ghana. BMC Health Serv Res 21, (2021).

14. Apeagyei, A. E. et al. Financing health in sub-Saharan Africa 1990–2050: Donor dependence and expected domestic health spending. PLOS Global Public Health 4, (2024).

15. Acharya, A. et al. The impact of health insurance schemes for the informal sector in low-and middle-income countries: A systematic review. World Bank Research Observer 28, 236–266 (2013).

16. Ochieng’ Odongo, S. SATISFACTION WITH THE QUALITY OF HEALTHCARE DELIVERY AMONG CLIENTS USING HEALTH INSURANCE IN NYERI COUNTY, KENYA. (2022).

17. Abraham, E. et al. Barriers and facilitators to health insurance enrolment among people working in the informal sector in Morogoro, Tanzania. Open Research Africa 4, 45 (2021).

18. Mulupi, S., Kirigia, D. & Chuma, J. Community Perceptions of Health Insurance and Their Preferred Design Features: Implications for the Design of Universal Health Coverage Reforms in Kenya. http://www.biomedcentral.com/1472-6963/13/474 (2013).

19. Dalinjong, P. A. & Laar, A. S. The national health insurance scheme: Perceptions and experiences of health care providers and clients in two districts of Ghana. Health Econ Rev 2, 1–13 (2012).

20. Bayked, E. M. et al. Willingness to pay for National Health Insurance Services and Associated Factors in Africa and Asia: a systematic review and meta-analysis. Frontiers in Public Health vol. 12 Preprint at 10.3389/fpubh.2024.1390937 (2024).

21. Mohammed, S., Sambo, M. N. & Dong, H. Understanding client satisfaction with a health insurance scheme in Nigeria: Factors and enrollees experiences. Health Res Policy Syst 9, (2011).

22. Kotoh, A. M., Aryeetey, G. C. & Van Der Geest, S. Factors that influence enrolment and retention in Ghana’ national health insurance scheme. Int J Health Policy Manag 7, 443–454 (2018).

23. Maritim, B. et al. “It is like an umbrella covering you, yet it does not protect you from the rain”: a mixed methods study of insurance affordability, coverage, and financial protection in rural western Kenya. Int J Equity Health 22, (2023).

24. Gatome-Munyua, A., Sieleunou, I., Sory, O. & Cashin, C. Why Is Strategic Purchasing Critical for Universal Health Coverage in Sub-Saharan Africa? Health Syst Reform 8, (2022).

25. Muhula, S., Opanga, Y., Kassim, S., Odeny, L., Mbewe, R. Z., Akoth, B., Jerop, M., Nyawira, L., Gueye, I., Kiplimo, R., Salamba, T., Kiarie, J., & Kimathi, G. (2025). Predictors of Health-Workforce Job Satisfaction in Primary Care Settings: Insights from a Cross-Sectional Multi-Country Study in Eight African Countries. International Journal of Environmental Research and Public Health, 22(7), 1108. 10.3390/ijerph22071108

26. Durizzo, K. et al. Toward mandatory health insurance in low-income countries? An analysis of claims data in Tanzania. Health Economics (United Kingdom) 31, 2187–2207 (2022).

27. Ekubu Otim, M. et al. Successes and Obstacles in Implementing Social Health Insurance in Developing and Middle-Income Countries: A Scoping Review of f-Year Recent Literatures. https://worldpopulationreview.com/country-.

28. Towards Sustainability Of The Community-Based Health Insurance In Rwanda: Successes, Challenges, And Opportunities | Strategic Purchasing Africa Resource Centre (SPARC). https://sparc.africa/2021/09/towards-sustainability-of-the-community-based-health-insurance-in-rwanda-successes-challenges-and-opportunities/.

29. Ly, M. S., Bassoum, O. & Faye, A. Universal health insurance in Africa: A narrative review of the literature on institutional models. BMJ Global Health vol. 7 Preprint at 10.1136/bmjgh-2021-008219 (2022).

30. Mathauer, I., Saksena, P. & Kutzin, J. Pooling arrangements in health financing systems: a proposed classification. doi:10.1186/s12939-019-1088-x.

31. Lutinah, R. R. Determinants of health insurance uptake in Tanzania

32. Mulupi, S., Kirigia, D. & Chuma, J. Community Perceptions of Health Insurance and Their Preferred Design Features: Implications for the Design of Universal Health Coverage Reforms in Kenya. http://www.biomedcentral.com/1472-6963/13/474 (2013).

33. Jehu-Appiah, C. et al. Equity aspects of the National Health Insurance Scheme in Ghana: Who is enrolling, who is not and why? Soc Sci Med 72, 157–165 (2011).

34. Moyehodie, Y. A., Mulugeta, S. S. & Yilema, S. A. The effects of individual and community-level factors on community-based health insurance enrollment of households in Ethiopia. PLoS One 17, e0275896 (2022).

35. Ziegler, S. et al. A step closer towards achieving universal health coverage: the role of gender in enrolment in health insurance in India. BMC Health Serv Res 24, (2024).

36. Zegeye, B. et al. Association between women’s household decision-making autonomy and health insurance enrollment in sub-saharan Africa. BMC Public Health 23, (2023).

37. Saksena, P., Antunes, A. F., Xu, K., Musango, L. & Carrin, G. Mutual health insurance in Rwanda: Evidence on access to care and financial risk protection. Health Policy (New York) 99, 203–209 (2011).

38. Uzochukwu, B. S. C. et al. Health care financing in Nigeria: Implications for achieving universal health coverage. Niger J Clin Pract 18, 437–444 (2015).

39. Ndayishimiye, C., Nduwayezu, R., Sowada, C. & Dubas-Jakóbczyk, K. Performance-based financing in Rwanda: a qualitative analysis of healthcare provider perspectives. BMC Health Serv Res 25, 1–12 (2025).

40. Sieleunou, I. et al. Setting performance-based financing in the health sector agenda: A case study in Cameroon. Global Health 13, 1–15 (2017).

